# Automated Extraction and Classification of Drug Prescriptions in Electronic Health Records: Introducing the PRESNER Pipeline

**DOI:** 10.1101/2023.10.04.23296481

**Authors:** Cristóbal Colón-Ruiz, Tomas Fitzgerald, Isabel Segura-Bedmar, Ewan Birney, Maria Herrero-Zazo

## Abstract

Electronic health record (EHR) systems with prescription data offer vast potential in pharmacoepidemiology and pharmacogenomics. The large amount of clinical data recorded in these systems requires automatic processing to extract relevant information. This paper introduces PRESNER, a name entity recognition (NER) and classification pipeline for EHR prescription data.

The pipeline uses the pre-trained transformer Bio-ClinicalBERT fine-tuned on UK Biobank prescription entries manually annotated with medication-related information (drug name, route of administration, pharmaceutical form, strength, and dosage) as the core NER system. Moreover, PRESNER also maps drugs to the Anatomical Therapeutic and Chemical (ATC) classification system and distinguishes between systemic and non-systemic drug products. It outperformed a baseline model combining the state-of-the-art Med7 and a dictionary-based approach from the ChEMBL database with a macro-average F1-score of 0.95 vs 0.71. In addition to UK Biobank prescription data, PRESNER can also be applied to other English prescription datasets, making it a versatile tool for researchers in the field.

## 1 Introduction

Large anonymized electronic health records (EHR) with prescription data linked to biobank registries enable large pharmacoepidemiologic and pharmacogenomic studies (Mcinnes, 2021). UK Biobank (UKB) collects health information and biological samples from over half a million volunteers (Bycroft *et al*., 2018). Since 2019, primary care records from the UK National Health System (NHS) have been linked for approximately 45% of the UKB cohort (∼230,000 participants), corresponding to more than 57 million prescription entries.

Unlike other prescription records (Pottegård *et al*., 2016; Alvarez-Madrazo *et al*., 2016), these datasets store unprocessed prescription information for dispensation purposes. Registries typically use national or internal codes for drugs, necessitating manual extraction by active ingredients and brand names (Fabbri *et al*., 2021).

An alternative to coding systems is to extract the information of interest from the text. This strategy also enables the extraction of additional information. In pharmacoepidemiology and pharmacogenomics using EHR data, a key need is identifying and grouping active ingredients, including synonyms and brand names. The Anatomical Therapeutic and Chemical (ATC) classification by WHO organises substances based on effects and properties, aiding international drug research (WHO, 2018). Additionally, the pharmaceutical form and route of administration must be considered to distinguish between drug products with systemic and non-systemic effects (e.g., oral vs topical antibiotics), while strength and dosage information are crucial for quantitative studies (Rodgers *et al*., 2017; Chen *et al*., 2018).

There has been substantial interest in developing Natural Language Processing (NLP) information extraction (IE) systems for drug named entity recognition (NER) (Segura-Bedmar *et al*., 2013; Kormilitzin *et al*., 2021) and other relevant medication information (Johnson *et al*., 2016). The irruption of contextual word embedding models, such as the Bidirectional Encoder Representations from Transformers (BERT) (Devlin *et al*., 2019), has fuelled the progress of NLP in all domains through the use of transfer learning with pre-trained models, especially when fine-tuned for a specific NLP application (Henry *et al*., 2020; Alsentzer *et al*., 2019).

This paper introduces PRESNER, a tailored NER and classification pipeline for EHR prescription data. Based on the transformer for medical text Bio-ClinicalBERT (Alsentzer *et al*., 2019) and fine-tuned with UKB prescription entries, PRESNER aims to automate standardised extraction, mapping, and classification of prescriptions, serving as a benchmarking tool for research acceleration. Anticipated broader applications include processing prescription data from sources like CPRD (Ghosh *et al*., 2019), MIMIC-III, and hospital EHR datasets.

PRESNER is designed to assist researchers working with raw prescription data from EHR. This pipeline not only performs drug NER but also offers additional capabilities, enabling users to automatically identify systemic drug products and apply filters to prescriptions based on drug name or therapeutic group by their ATC codes. The workflow seamlessly handles entity recognition and classification tasks, by mapping prescriptions to ATC codes and categorising them as either systemic or non-systemic. Users can choose to use the post-processing output or access the results of the NER component to tailor solutions to their specific research questions. The pipeline is available at https://github.com/ccolonruiz/PRESNER.

## 2. Materials and Methods

### 2.1 ChEMBL dictionary

The ChEMBL database (Gaulton *et al*., 2012) is the largest open-source resource for medicinal chemistry data. Using its web services API, we built a dictionary for approved human molecules. The latest version of the dictionary contained 38,215 synonyms for 4,127 active ingredients, mapped to 4,193 ChEMBL ID and 3,206 ATC codes. However, to include the latest information curated from the database, when run by the user the pipeline accesses the API to create an updated version of the dictionary. This dictionary is used by the pipeline to map prescription entries to their ATC codes. We also used this dictionary in a baseline approach for the NER task.

### 2.2 UK Biobank prescription dataset

UKB Data-Field “42039” was used to retrieve prescription data, covering primary care prescriptions for 222,096 individuals and 102,052 unique prescription names entries. These data included the drug product’s name, quantity, prescription date, and one or more codes from medicinal and sanitary products coding systems used across the NHS primary care (Supplementary Table 1).

A dataset of 1,100 randomly selected prescription entries for fine-tuning and evaluation was manually annotated using the BRAT tool (Peng *et al*., 2019) by a pharmacist with prior experience in the annotation of pharmacological texts (Table 1). We applied stratified sampling to split the whole dataset into train (80%) and test (20%) splits. We refer to this annotated dataset as the UKB prescription dataset throughout this document.

**Table 1.** Distribution of UK Biobank (UKB) and n2c2 annotated text spans used for training and test. The document types are prescription entries and discharge summary entries in the UKB prescription dataset and the n2c2 corpus, respectively.

|  | UKB prescription dataset |  | n2c2 corpus |  |
| --- | --- | --- | --- | --- |
|  | Training (80%) | Test (20%) | Training (60%) | Test (40%) |
| Dosage | 822 | 206 | 4,227 | 2,681 |
| Drug | 817 | 206 | 16,257 | 10,551 |
| Form | 1168 | 302 | 6,657 | 4,359 |
| Route | 92 | 27 | 5,460 | 3,513 |
| Strength | 884 | 215 | 6,694 | 4,230 |
| Total documents | 880 | 220 | 303 | 202 |

### 2.3 The n2c2 corpus

Due to annotation processes being one of the most time-costly and expensive tasks in NLP and machine learning, the size of our UK Biobank prescription dataset is limited. For this reason, we also exploited the National NLP Clinical Challenges (n2c2) corpus to fine-tune our NER pipeline. This corpus includes over 83 thousand annotated entities for medication information (strength, dosage, duration, frequency, form, route, reason for administration, and adverse effects) from 505 discharge summaries (Table 1) from the MIMIC-III dataset (Johnson *et al*., 2016).

### 2.4 The NER task

The core model in PRESNER is based on Bio-ClinicalBERT (Alsentzer *et al*., 2019), a contextual word embedding model that builds upon BioBERT (Lee *et al*., 2020) and specifically pre-trained on the MIMIC-III dataset, addressing the linguistic disparities between general domain and clinical texts.

We fine-tuned the Bio-ClincalBERT model by adding a conditional random field (CRF) layer (Ketmaneechairat and Maliyaem, 2020). This layer allows us to capture dependencies between labels. The extended architecture – i.e., the Bio-ClinicalBERT model enhanced with a CRF optimization layer (Supplementary Figure 1) – will be referred to as the BCB-CRF model throughout this document. Pre-processing and training details are provided in Supplementary Methods.

For comparison purposes, the BCB-CRF model was fine-tuned in the following three scenarios: i) only using the n2c2 corpus, ii) only using the UKB prescription dataset, and iii) using both datasets. In this third scenario (Figure 1), the BCB-CRF model was fine-tuned on the n2c2 corpus, and then, a second fine-tuning was performed on the UKB prescription dataset. The n2c2 corpus was used first due to the relatively limited number of annotated entities in the UKB prescription dataset (Table 1). Moreover, this approach allows us to harness the knowledge transfer from the n2c2 corpus and also adapt to the unique language features present in the UKB prescription dataset. Our hypothesis is that this approach can improve the recognition of under-represented entities such as ROUTE (with only 92 instances in the training split of the UKB prescription dataset). For hyperparameter tuning, 20% of the n2c2 training sentences were used as a validation set. Limited annotated data in the UKB prescription dataset necessitated 5-fold cross-validation.

**Figure 1.**
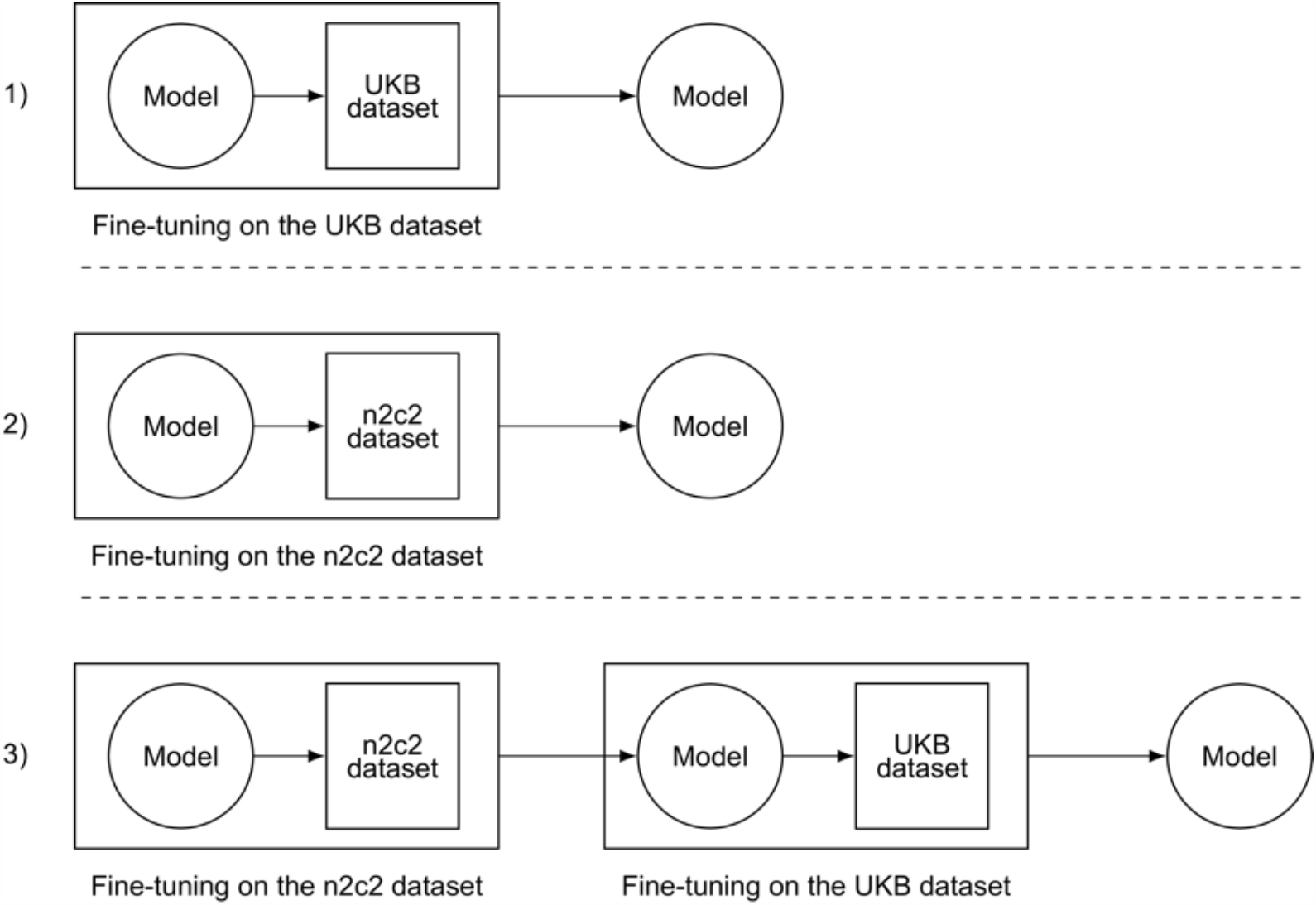
Representation of the fine-tuning approaches compared in this work. 1) Fine-tuning on the UKB prescription dataset. 2) Fine-tuning on the n2c2 corpus. 3) Fine-tuning on the n2c2 corpus and UKB prescription dataset.

We also developed a baseline approach combining a dictionary-based NER module based on the ChEMBL dictionary and the pre-trained NER models from Med7 (Kormilitzin *et al*., 2021). There are two Med7 models available, one based on convolution networks and another based on the transformer-based model RoBERTA (Liu *et al*., 2021). In this work, we combined the output of both and refer to them as Med7 for simplicity. The output of the baseline is the union of all entities identified by the dictionary and Med7. More details on the baseline model can be found in Supplementary Methods.

### 2.5 Classification tasks: mapping ATC codes and detecting drug products with systemic activity

After the NER phase, PRESNER aims to classify drug products based on the extracted information in the previous NER step, differentiating between those with and without systemic activity and assigning the most appropriate ATC code(s). This facilitates further filtering by researchers using the EHR prescription dataset.

ATC codes were mapped to DRUG entities using a refined dictionary created from ChEMBL (including 3,206 ATC codes linked to 3,433 ChEMBL ID). Moreover, the ROUTE and FORM entities annotated in the previous phase (see section 2.4) were manually reviewed to create dictionaries for systemic and non-systemic administration routes and pharmaceutical forms. These dictionaries for pharmaceutical form and administration route are used to classify prescriptions as systemic (SDP) or non-systemic drug products (NSD) and to select the preferred ATC codes (Supplementary Figure 2).

## 3 Results

The baseline approach was outperformed by the BCB-CRF model which was either fine-tuned using only the UKB prescription dataset or initially fine-tuned with n2c2 followed by fine-tuning with the UKB prescription dataset. These two BCB-CRF models obtained higher F1 for all entity types (Table 2). The greatest improvement (up to 0.24 in macro-F1) was achieved by the BCB-CRF model fine-tuned first on the n2c2 corpus and then on the UKB prescription dataset. While transfer learning from n2c2 corpus helped to mitigate the scarcity of annotated entities in the UKB prescription dataset, this second dataset provided information on the text distribution of the target documents (i.e., prescription entries). The effectiveness of this approach was particularly evident in the case of the under-represented entity type, ROUTE, where pre-adjustment with n2c2 significantly improved recall performance.

**Table 2.**
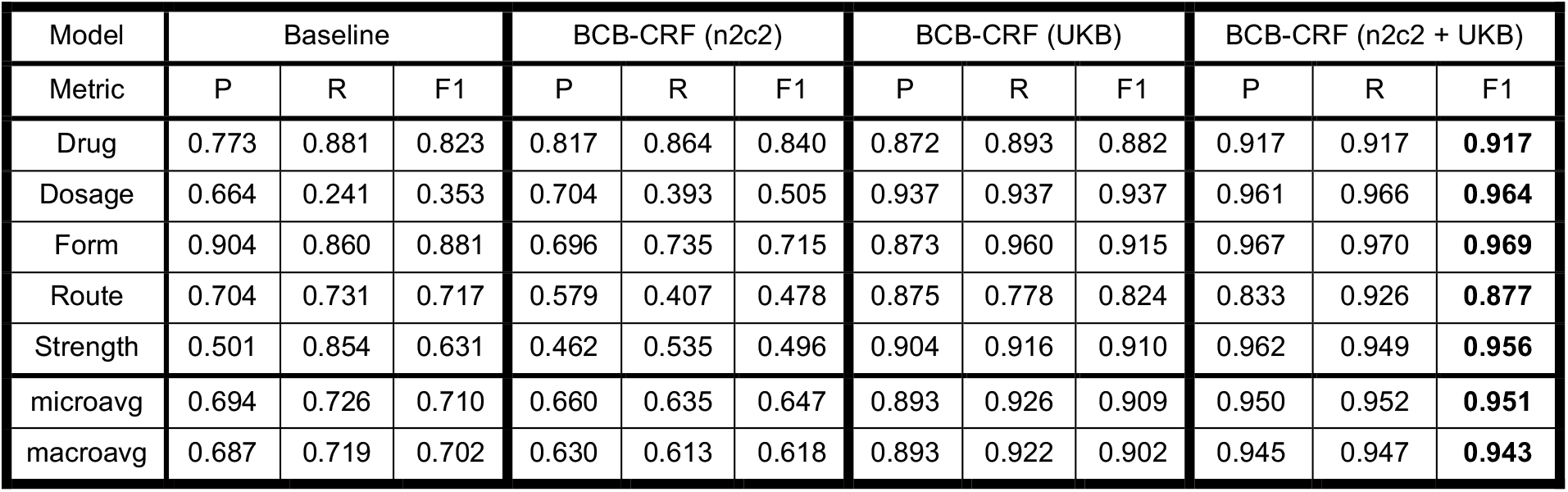
Comparison of results of the baseline and three BCB-CRF models on the hold-out test showing precision (P), recall (R) and F1-score (F1). BCB-CRF (n2c2): model fine-tuned with the n2c2 corpus. BCB-CRF (UKB): model fine-tuned with the UKB prescription dataset. BCB-CRF (n2c2 + UKB): model fine-tuned with the n2c2 corpus and the UKB prescription dataset.

| Model | Baseline |  |  | BCB-CRF (n2c2) |  |  | BCB-CRF (UKB) |  |  | BCB-CRF (n2c2 + UKB) |  |  |
| --- | --- | --- | --- | --- | --- | --- | --- | --- | --- | --- | --- | --- |
| Metric | P | R | F1 | P | R | F1 | P | R | F1 | P | R | F1 |
| Drug | 0.773 | 0.881 | 0.823 | 0.817 | 0.864 | 0.840 | 0.872 | 0.893 | 0.882 | 0.917 | 0.917 | **0.917** |
| Dosage | 0.664 | 0.241 | 0.353 | 0.704 | 0.393 | 0.505 | 0.937 | 0.937 | 0.937 | 0.961 | 0.966 | **0.964** |
| Form | 0.904 | 0.860 | 0.881 | 0.696 | 0.735 | 0.715 | 0.873 | 0.960 | 0.915 | 0.967 | 0.970 | **0.969** |
| Route | 0.704 | 0.731 | 0.717 | 0.579 | 0.407 | 0.478 | 0.875 | 0.778 | 0.824 | 0.833 | 0.926 | **0.877** |
| Strength | 0.501 | 0.854 | 0.631 | 0.462 | 0.535 | 0.496 | 0.904 | 0.916 | 0.910 | 0.962 | 0.949 | **0.956** |
| microavg | 0.694 | 0.726 | 0.710 | 0.660 | 0.635 | 0.647 | 0.893 | 0.926 | 0.909 | 0.950 | 0.952 | **0.951** |
| macroavg | 0.687 | 0.719 | 0.702 | 0.630 | 0.613 | 0.618 | 0.893 | 0.922 | 0.902 | 0.945 | 0.947 | **0.943** |

On the contrary, the BCB-CRF model fine-tuned only on the n2c2 corpus provided lower results than the other BCB-CRF models and the baseline approach for FORM, ROUTE, and STRENGTH entities. In addition to the disparities in text distributions between n2c2 and prescription entries, this lower performance could be due to the fact that Med7 was trained with n2c2 but also with two additional corpora annotated by the authors.

The PRESNER pipeline also addresses the mapping of DRUG entities to ATC codes by using a matching process based on a dictionary from ChEMBL. DRUG entities extracted by the BCB-CRF or the Med7 module of the baseline but not mapped to ATC codes were mainly brand names not included in the ChEMBL dictionary (Figure 2 and Supplementary Figures 3 and 4), like hormone replacement therapy or oral contraceptives, and non-drug items, supplements, or healthcare terms. A word cloud (Figure 2) of top terms not retrieved by any approach (10% total) displays non-medicinal sanitary products predominantly.

**Figure 2.**
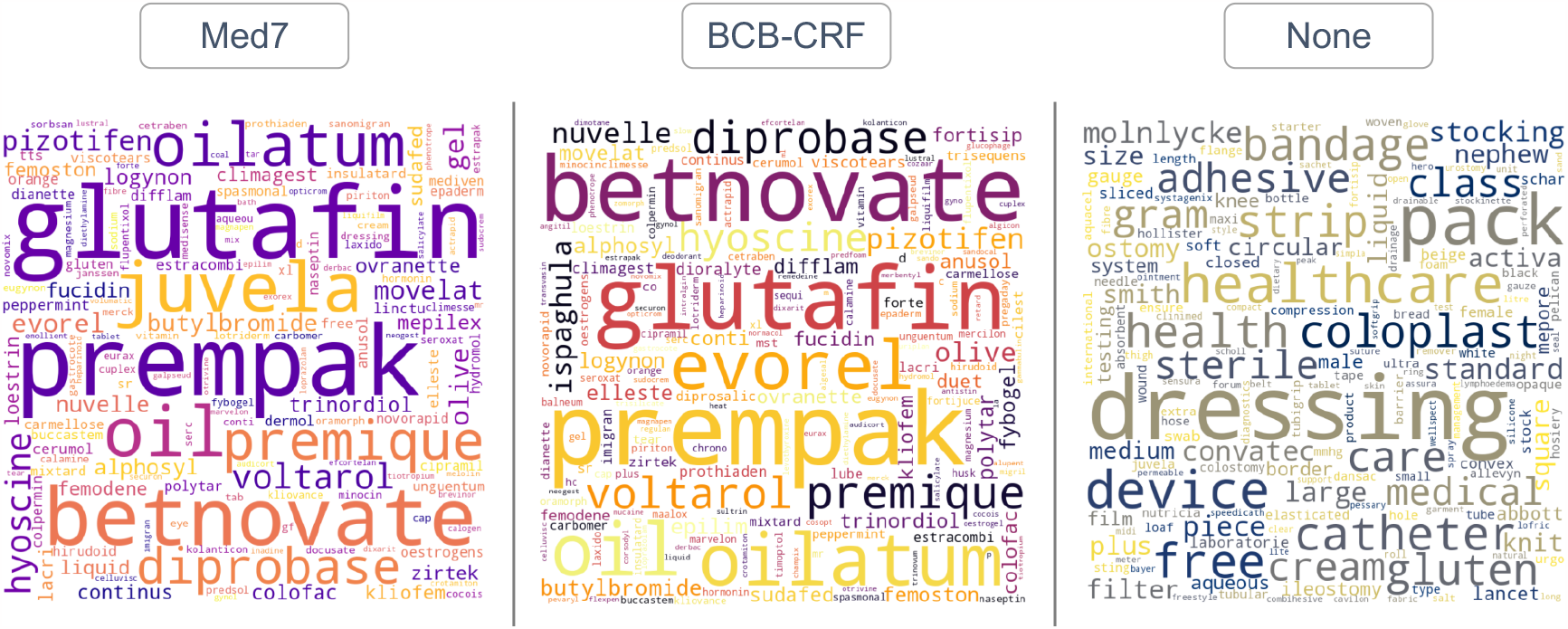
Word-clouds of top entities retrieved but not mapped to ATC codes by only one of the models: the MED7 module of the baseline model or the BCB-CRF model. The third panel shows the top terms from drug product entries with no entity DRUG retrieved by any of the drug NER models.

**Figure 3.**
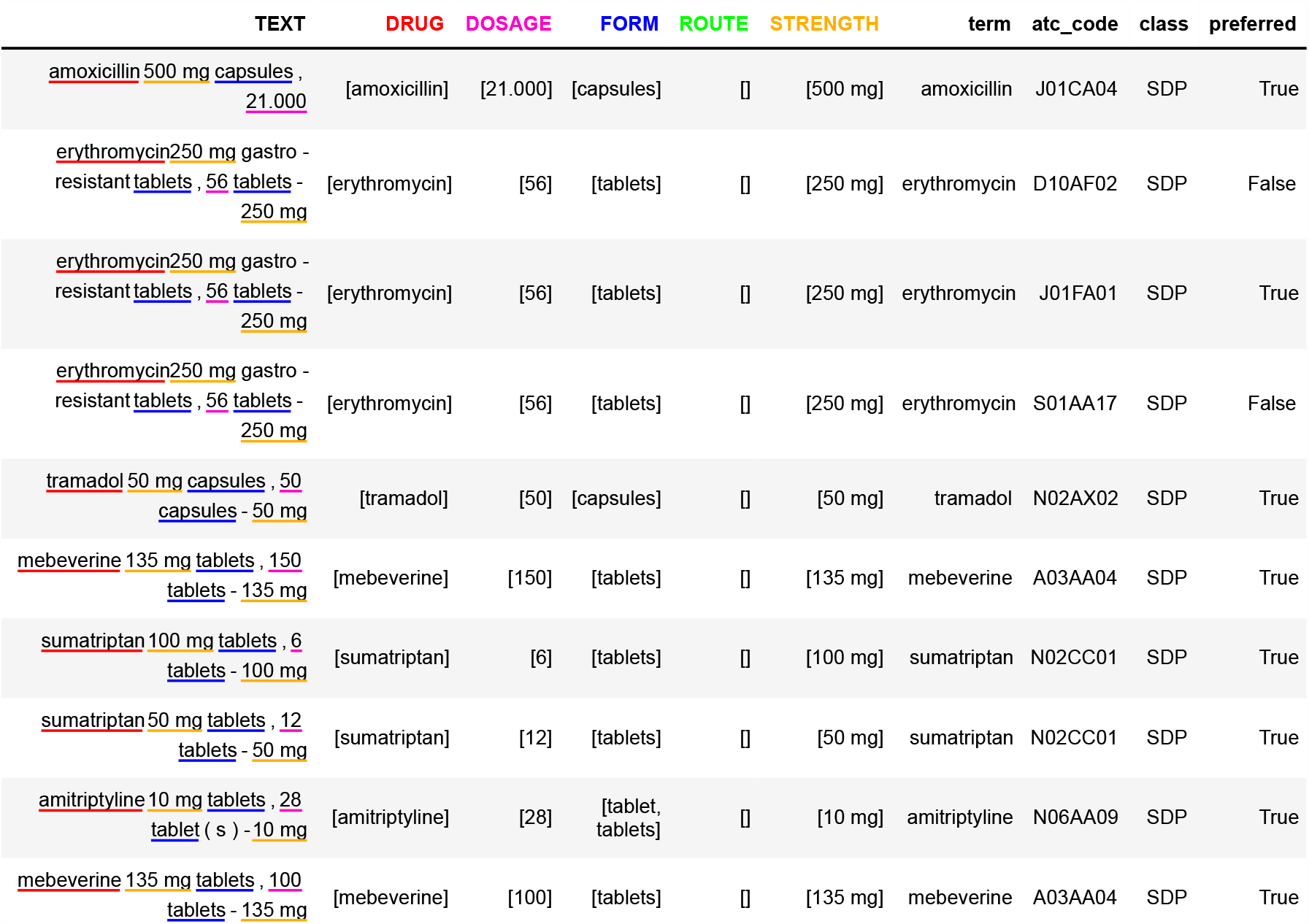
Example of drug information extracted by PRESNER. *TEXT*: text of the prescription entry with the entity types highlighted in different colours.; *DRUG, DOSAGE, FORM, ROUTE, STRENGTH*: extracted entities for each type; *term*: drug name in ChEMBL dictionary mapped to the DRUG entity; *atc code*: mapped ATC codes for that term; *class*: drug product classification as systemic (SDP), non-systemic (NSDP) or potential systemic drug product (PSDP); *preferred*: label for the ATC code as preferred or non-preferred based on the assigned class.

**Figure 4.**
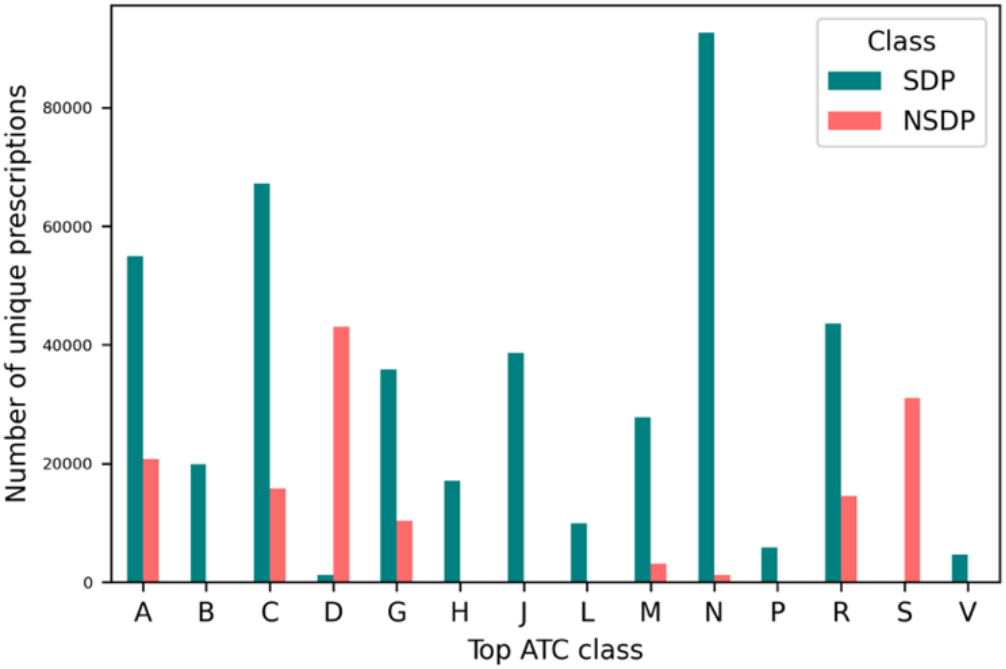
Number of unique prescription entries by top ATC class classified as systemic drug product (SDP) or non-systemic drug product (NSDP). One prescription can be mapped to more than one ATC codes and categories.

The baseline retrieved more incorrect ROUTE terms (e.g., the pharmaceutical form ‘injection’, or the adjectives ‘dispersible’, ‘gastro’, ‘conjugated’) while failing more often to retrieve correct ones such as ‘eye’, ‘ear’, ‘vaginal’ or ‘oromucosal’. All the approaches failed to extract correct FORM terms, but non-related words (e.g., ‘medicated’, ‘pack’, ‘‘vanilla’, ‘lasagne’, ‘blister’) were more commonly retrieved by Med7 (Supplementary Figure 5).

Fine-tuning with both n2c2 and the UKB prescription dataset greatly benefited the entities STRENGTH and DOSAGE compared to the baseline. The baseline model performed poorly for entity DOSAGE with precision and recall scores of 0.66 and 0.24, respectively. Likewise, entity STRENGTH had a precision of only 0.5, whereas the BCB-CRF model achieved a precision of 0.96. This is partly because the baseline consistently mislabelled the total prescribed unit count as STRENGTH instead of DOSAGE, resulting in low recall for DOSAGE (0.2) and low precision for STRENGTH (0.5).

Figure 3 shows an example of the output of the PRESNER pipeline, with the extracted structured information for the various entity types. 458,132 prescription entries (72% of the total dataset) were mapped to at least one ATC code. From these, 81% and 13% were classified as SDP and NSDP, respectively.

Preferred ATC codes were selected by the pipeline based on this classification, leading to the distribution of top ATC classes shown in Figure 4. Prescription entries that could not be clearly classified by PRESNER (6%) were labelled as potential SDP (PSDP).

## Discussion and Conclusions

Prescription data linked to large biobanks enable pharmacogenomic studies (McInnes *et al*., 2021), but processing these raw data is complex and time consuming. Manual lists of names and codes are frequently used for UKB primary care data (Darke *et al*., 2022; Fabbri *et al*., 2021; Topless *et al*., 2022). However, these terminologies have inherent flaws such as variations in NHS implementation across time and regions. (Spiers *et al*., 2017).

We demonstrate the superiority of our fine-tuned BCB-CRF model over a baseline combining the state-of-the-art Med7 and a dictionary-based approach. While dictionaries often exhibit high precision, they lack recall due to synonym diversity. Machine learning, especially transformer models, excel in capturing contextual cues for entity identification. This proficiency allows transformers to effectively address NLP ambiguity by recognizing context-based representations.

Pre-training on clinical documents enables BCB-CRF to capture relevant clinical knowledge and context. Fine-tuning with n2c2 and the annotated UKB prescription dataset further enhances performance by refining its representations and parameters to align with the specific task requirements. This combined approach optimally uses both pre-trained models and task-specific annotations for better clinical NLP tasks.

Finally, the regular structure of prescription entries in the UKB prescription dataset greatly favours the use of transfer learning that can substantially reduce the heavy workload of the annotation tasks.

PRESNER has demonstrated its effectiveness in extracting structured information from prescription entries, including drug names, strength, dosage, pharmaceutical form, and route of administration. The pipeline also enables classification tasks such as mapping drug names to the ATC classification system and classifying prescription entries as systemic or non-systemic based on extracted route and form information. Additionally, the extracted strength and dosage data can, for example, be utilised to infer the total prescribed dose for quantitative analysis. Moving forward, our future work aims to incorporate additional functionalities, such as identifying co-medications or cleaning and imputing missing strength or dosages.

It is important to acknowledge certain limitations. Some drugs may have not been recognised by the NER pipeline or mapped by the ChEMBL dictionary. After initial manual revision, 65 drug entities with a frequency greater than 100 in the current dataset but not included in the ChEMBL dictionary (e.g., ‘*co-codamol’*, ‘*valproate’*, or ‘*malarone’*) were added manually. Even so, drugs like insulins, hormone replacement therapies, contraceptives, and brand names for comedications may not have been consistently identified. Future enhancements could focus on improving the dictionaries or incorporating pattern-based rules to enhance the recall of the PRESNER pipeline. Although the pipeline discerns between systemic and non-systemic ATC codes, some molecules have more than one ATC code and it currently does not identify a singular ATC code for each drug product. Finally, manual validation of the extracted results for all drug products has not been done and, therefore, manual review by the users is encouraged.

Looking ahead, as the UK Biobank secures access to primary care data for the remaining 35% of the cohort, we anticipate that PRESNER will prove to be a valuable resource for processing these new and extensive datasets. Furthermore, the potential for utilizing the same pipeline to process other English prescription databases is promising, showcasing its potential for wider application in the field.

## Data Availability

UK Biobank data are available under restricted access through a procedure described at http://www.ukbiobank.ac.uk/using-the-resource/.
All other data produced in the present work and the code are available at https://github.com/ccolonruiz/PRESNER

## Availability

https://github.com/ccolonruiz/PRESNER

## Funding

This research was supported by the Novo Nordisk Foundation (NNF17OC0027594).

## Acknowledgments

We acknowledge ChEMBL for their advice on the creation of the dictionaries and use of the API.

## Notes

### Competing Interest Statement

The authors have declared no competing interest.

### Author Declarations

The study used available human data that were originally located at the UK Biobank resource.

