## Supplementary Methods for "Automated Extraction and Classification of Drug Prescriptions in Electronic Health Records: Introducing the PRESNER Pipeline"

### Pre-processing

The prescription entries were pre-processed by combining the drug name and quantity descriptions, transforming the texts into lowercase letters, and separating the values from units of measurement to homogenise their format (e.g., the drug name "*Erythromycin 250mg gastro-resistant tablets*" and quantity "*56 tablets - 250 mg*" results in the prescription entry "*erythromycin 250 mg gastro-resistant tablets , 56 tablets - 250 mg*".

The texts were tokenized using the pre-trained Bio-ClinicalBERT model vocabulary (28,996 tokens) and the tokenizer provided by the transformer's library.[1] Most texts range from 10 to 60 tokens based on the probability density function (Supplementary Methods Figure 1). Medical prescription texts, in particular, have a maximum length of 53 tokens, while sentences in the 2018 n2c2 dataset are longer than prescriptions. To include all examples from both datasets without data truncation, the text length was adjusted to 225 tokens – i.e., the length of the longest text in the dataset. This approach avoids information loss and ensures comprehensive coverage.

[1] https://huggingface.co/docs/transformers/index

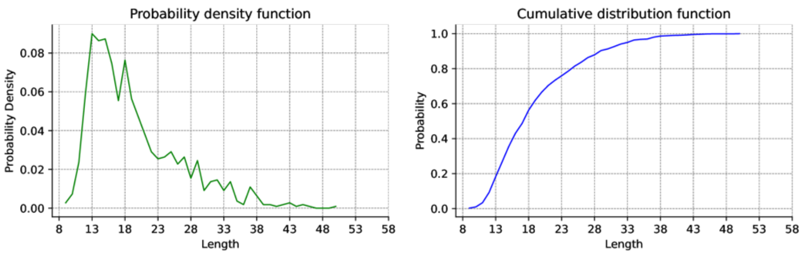


Probability density function and Cumulative distribution function over the length of prescriptions (UKB dataset).

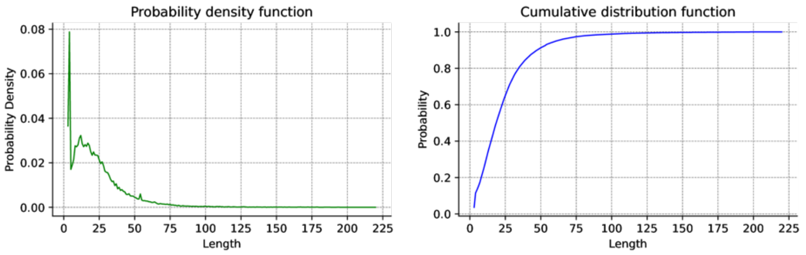


Probability density function and Cumulative distribution function over the length of EHR sentences (2018 n2c2 dataset).

Supplementary Methods Figure 1. Probability density function and Cumulative distribution function over the length of prescriptions in the UKB dataset and the n2c2 corpus.

### Baseline Model (ChEMBL dictionary and Med7)

The baseline approach combined a dictionary-based NER module based on the ChEMBL dictionary and the pre-trained NER model Med7. In the first case, drug name entities were extracted by matching texts with the term in the dictionary. The output of this approach was combined with the output of Med7. We refer to this as the baseline model.

Med7 is a NER model trained on clinical notes from both n2c2 and secondary care mental health EHR notes databases (UK-CRIS) to extract seven entity types: drug name, dosage, duration, form, frequency, route, and strength. There are two variants of the Med7 model: Med7_lg and Med7_trf. Med7_lg uses a bloom embeddings linguistic model (Serrà and Karatzoglou, 2017) pre-trained on the entire MIMIC-III dataset to initialise the weights of several residual convolutional layers. The convolutional layers provide contextual information about the words, which is used as a transition-biased approach to identify entities. On the other hand, Med7_trf employs the RoBERTa transformer-based linguistic model to generate word embedding with contextual information for the transition-based approach. In this research, the outputs of the two models were combined and referred to as the Med7 approach.

<u>The NER model in PRESNER</u>

Bio-ClinicalBERT (Alsentzer *et al.*, 2019) is a contextual word embedding model that builds upon BioBERT (Lee *et al.*, 2020) and was specifically trained on the MIMIC III clinical notes dataset, addressing the linguistic disparities between general domain texts and clinical texts. Bio-ClinicalBERT utilises 12 encoding layers with a hidden layer dimension of 768, resulting in approximately 110 million trainable parameters.

Contextual word embeddings from Bio-ClinicalBERT allows to capture dependencies within texts. The transformer is fine-tuned by adding a conditional random field (CRF) layer (Ketmaneechairat and Maliyaem, 2020) such as the linear-chain CRF, which offers the advantage of capturing dependencies between labels. This sequential dependency between label $yk$ and the previous label $yk-1$, as demonstrated in the equation below, can be effectively captured. Here, $\bar{x}$ represents a sequence of words, $\bar{y}$ represents the corresponding sequence of labels, and f represents a feature function that decomposes into $U(xk,yk)$ denoting the score assigned to the observation features, and $T(yk,yk+1)$ denoting the score assigned to the transition between consecutive labels $yk$ and $yk-1$. The equation also includes $Z(\bar{x})$, which represents the partition function (a normalising sum of all possible feature scores for all possible label sequences).

$$P(\bar{y}|\bar{x}) = \frac{\prod_{k=1}^{K} f(y_k, y_{k-1}, x_k)}{\sum_{y'} \prod_{k=1}^{K} f(y'_k, y'_{k-1}, x_k)} = \frac{\exp(\sum_{k=1}^{K} U(x_k, y_k) + \sum_{k=1}^{K} T(y_k, y_{k-1}))}{Z(\bar{x})}$$

To evaluate the performance of our model, we compared the performances of 1) the baseline model - i.e., ChEMBL-based approach combined with Med7; 2) the Bio-ClinicalBERT model with the CRF optimization layer (from now on referred to as the BCB-CRF model through this document) fine-tuned using only the UKB dataset; 3) the same model fine-tuned using the n2c2 training set followed by UKB data; 4) the same model fine-tuned with the n2c2 corpus only. For hyperparameter tuning, 20% of the n2c2 training sentences were used as a validation set. Limited annotated data in the UKB set necessitated 5-fold cross-validation. Both cases employed a dropout of 0.3, RMSprop optimizer with a learning rate of 3e-5, and a batch size of 128 for five epochs.
