## Supplementary Material for "Automated Extraction and Classification of Drug Prescriptions in Electronic Health Records: Introducing the PRESNER Pipeline"

**Supplementary Figure 1.** Architecture overview of the Bio-ClinicalBERT model with the CRF optimization layer (BCB-CRF)

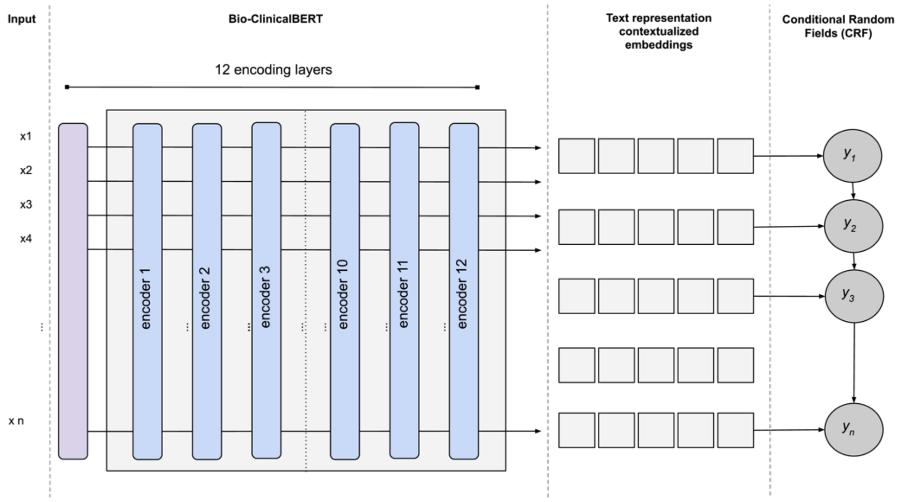


**Supplementary Figure 2.** Diagram of rule-based approach to classify prescription entries as non-systemic (NSDP), systemic (SDP), or potentially systemic (PSDP) drug products.

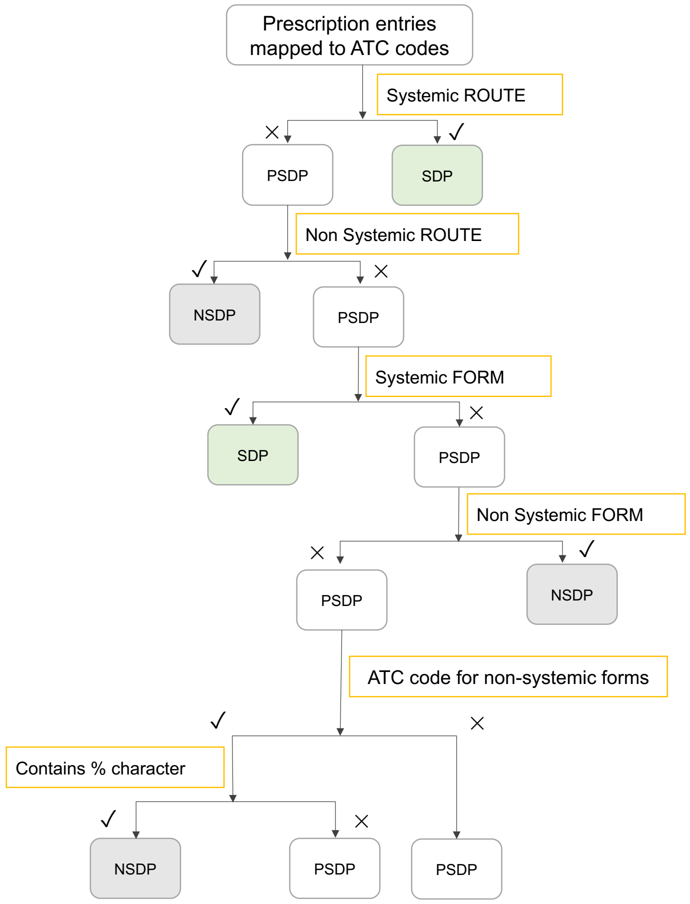

**Supplementary Figure 3.** Percentage of prescription entries processed by three different Drug NER approaches. The three categories displayed are prescription with at least one drug name retrieved and mapped to ATC code, prescriptions retrieved but not mapped to ATC codes, and prescriptions with none retrieved DRUG entity.

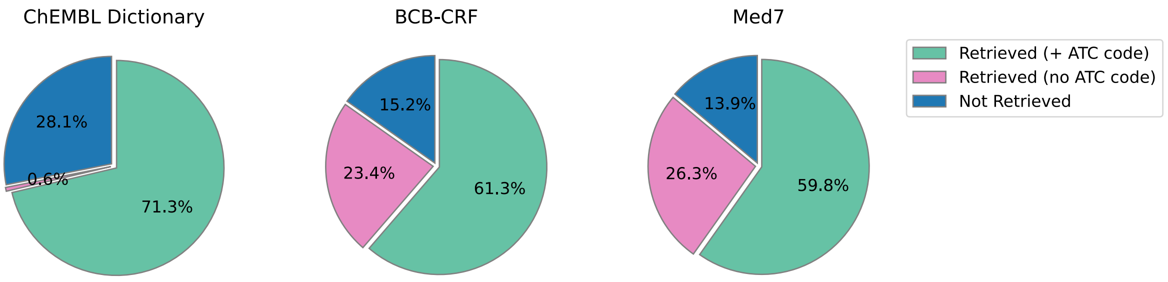


**Supplementary Figure 4.** Overlap of prescription entries where drug names were successfully retrieved and mapped to ATC codes by the three drug NER approaches: the ChEMBL dictionary and Med7 mudules in the baseline and the BCB-CRF model.

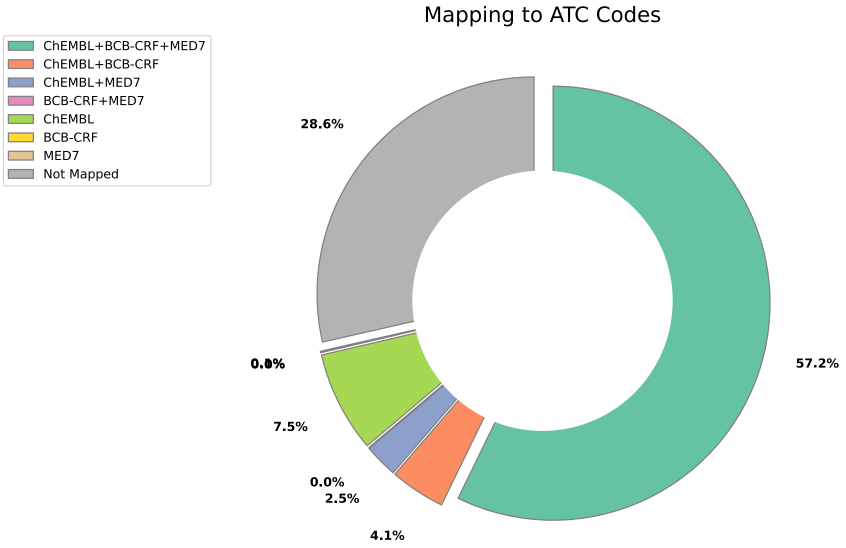

**Supplementary Figure 5.** Word-clouds of top entities ROUTE and FORM retrieved only by the Med7 module of the baseline model or only by the BCB-CRF model.

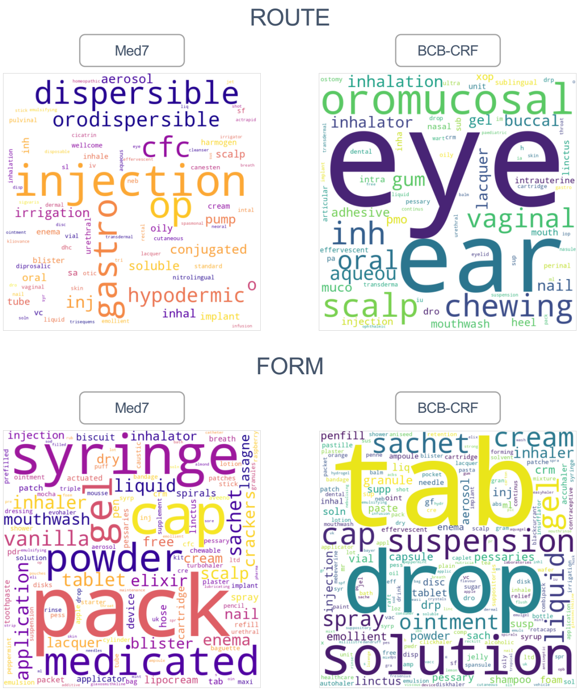


**Supplementary table 1.** Number of prescription entries mapped to the various NHS coding systems in the UKB primary care prescription dataset.

| Coding system | Number of unique prescription name |
|---|---|
| BNF[1] | 41942 |
| dm+d[2] | 12672 |
| Read v2[3] | 2 |
| Read v2 & dm+d | 39953 |
| Read v2 & BNF | 6135 |
| BNF v2 & dm+d | 0 |
| No code | 1348 |

1. The British National Formulary (BNF) is the standard list of medicines and sanitary products prescribed in the UK; 2. The Dictionary of Medicines and Devices (dm+d) is a dictionary of descriptions and codes that represent medicines and devices in use across the NHS, available for the EHR primary practice management system Vision in England (but not by the other practice management systems or the Vision versions used in Scotland or Wales). 3. Read codes are a coded thesaurus of clinical terms used in primary care since 1985 which covers drug products and sanitary products. The different versions of Read Codes are now deprecated, and SNOMED CT has been introduced in the NHS system. Although SNOMED CT has not been included in the current version of UK Biobank, it should be expected that it will in further releases.
